# An Annotated Multi-Site and Multi-Contrast Magnetic Resonance Imaging Dataset for the study of the Human Tongue Musculature

**DOI:** 10.1101/2024.12.09.24318591

**Authors:** Fernanda L. Ribeiro, Xiangyun Zhu, Xincheng Ye, Sicong Tu, Shyuan T. Ngo, Robert D. Henderson, Frederik J. Steyn, Matthew C. Kiernan, Markus Barth, Steffen Bollmann, Thomas B. Shaw

## Abstract

This dataset provides the first annotated, openly available MRI-based imaging dataset for investigations of tongue musculature, including multi-contrast and multi-site MRI data from non-disease participants. The present dataset includes 47 participants collated from three studies: BeLong (four participants; T2-weighted images), EATT4MND (19 participants; T2-weighted images), and BMC (24 participants; T1-weighted images). We provide manually corrected segmentations of five key tongue muscles: the superior longitudinal, combined transverse/vertical, genioglossus, and inferior longitudinal muscles. Other phenotypic measures, including age, sex, weight, height, and tongue muscle volume, are also available for use. This dataset will benefit researchers across domains interested in the structure and function of the tongue in health and disease. For instance, researchers can use this data to train new machine learning models for tongue segmentation, which can be leveraged for segmentation and tracking of different tongue muscles engaged in speech formation in health and disease. Altogether, this dataset provides the means to the scientific community for investigation of the intricate tongue musculature and its role in physiological processes and speech production.

## Background and Summary

The human tongue is involved in many physiological processes^1,2^ – including food manipulation and recognition^1,3^, taste^2^, thermosensation^4,5^, and breathing^6^ – and speech^7,8^. This diverse set of functionalities characterises the tongue as both a motor and a sensory organ^2^. Moreover, to subserve these processes, different tongue muscles may compress and/or elongate, but the overall tissue volume is constant, i.e., the tongue is a muscular hydrostat^6^. The tongue is also implicated in neurodegenerative diseases^9,10^, developmental speech pathologies^11^, and sleep disorders^12^, making it a potential biomarker for conditions affecting function. For example, studies have found diffuse T1-weighted hyperintensity of the tongue musculature in Amyotrophic Lateral Sclerosis (ALS) patients^13,14^ and reduced tongue volume in ALS patients with bulbar palsy^15^. Despite the potential of the measures of the tongue (e.g., morphometry and volume) as a biomarker, there exists no comprehensive annotated and publicly available MRI dataset to help inform understanding of tongue anatomy in health and disease. We, therefore, introduce this critical resource that will enable the identification of new biomarkers and interventions.

The tongue is comprised of anatomically distinguishable and interconnected intrinsic and extrinsic muscles^16,17^. The intrinsic muscles of the tongue both originate and insert within the tongue itself. There are four pairs of these intrinsic muscles: the superior longitudinal, inferior longitudinal, transverse, and vertical muscles. In contrast, the extrinsic muscles originate from structures outside the tongue, including the genioglossus, hyoglossus, styloglossus, and palatoglossus muscles. Each of these muscles can move the tongue in a particular direction^6^, but coordinated contractions of multiple muscles work together to enable movements like protrusion, retraction, and elevation of the tongue, as well as changing its shape and position during activities such as chewing and swallowing^18^.

Magnetic resonance imaging (MRI) is an important non-invasive technique that allows imaging and identification of the intricate tongue musculature^19^. Accordingly, in our recent study^20^, we compiled a detailed guideline for the identification and segmentation of the superior longitudinal, transverse/vertical combined, genioglossus and inferior longitudinal muscles of the tongue. Although detailed manual annotation was performed using T2-weighted (T2w) images – mostly from patients with motor neuron disease (MND) – where the contrast differences were clear across these different muscles, we developed a semi-automated segmentation pipeline for tongue segmentation of participants across different studies and MRI contrasts [T1-weighted (T1w) and T2-weighted images]. Here, we provide a subset of that data, i.e., the data from non-disease controls, to aid future research into tongue anatomy.

This is the first openly available fully annotated MRI-based imaging data for tongue segmentation^21^. This dataset includes structural MRI data (T2w and/or T1w) from non-disease controls, with accompanying template space and demographic information, from three studies/scanners. We provide automatically generated and manually corrected tongue segmentation labels to facilitate further research requiring annotated data. Additionally, we distribute an atlas generated from these manually corrected segmentations to provide a more accurate model of tongue muscle location and size. This dataset can serve as a valuable resource for developing and refining machine learning models for tongue segmentation, which, in turn, can enhance automated analysis of tongue movement in speech production using high-speed real-time MRI (e.g., CINE)^22,23^. Similarly, these data can be used to inform longitudinal studies of disease progression, particularly in conditions where tongue function is affected, e.g., bulbar onset ALS^24^. Given that the tongue is routinely captured in standard MRI scans of the brain and head/neck, the annotated data and atlas provided here can also serve as a reference for analysing existing and future MRI datasets, broadening its utility across multiple research domains. In sum, this new imaging/segmentation dataset of the human tongue provides the means for the scientific community to investigate tongue musculature and its role in physiological processes and speech production.

## Methods

### Participants

The present dataset^21^ includes information from 47 non-neurodegenerative “healthy” participants (20 females, 25-80 years old) collated from three studies: The Biomarkers of Long surviving MND (BeLong; 4 participants), Exploring Appetite Targets and Therapies for Motor Neuron Disease (EATT4MND;19 participants), and the Brain and Mind Centre Motor Neuron Disease neuroimaging database (BMC; 24 participants). Table 1 shows basic demographic information across studies. Note, however, that the original studies include data from patients, which we chose not to include on our public data release for three reasons: 1) Ethical considerations: releasing patient data openly is subject to strict privacy regulations; 2) Data quality: we prioritised quality over quantity, as all segmentations in our public release were manually curated. Patient scans often suffer from motion artifacts due to increased movement and decreased SNR, making it more challenging to objectively define muscle boundaries; 3) Demonstrated generalisability: in a separate study, we have shown that models trained on this dataset can successfully segment tongues in patients, even when significant morphological and volumetric changes are present^20^.

**Table 1.** Demographic information.

| ***Study*** | ***Number of individuals***<br>***(number of females)*** | ***Age range (years)*** | ***Weight/Height***<br>***availability*** |
| --- | --- | --- | --- |
| ***BeLong*** | 4 (2) | 25-74 | Available |
| ***EATT4MND*** | 19 (5) | 26-73 | Available |
| ***BMC*** | 24 (13) | 37-80 | Not available |

### Ethics

All studies were approved by their relevant Human Research Ethics Committees. Specifically, BeLong was approved by the University of Queensland HREC (2021/HE000975), EATT4MND was approved by the University of Queensland HREC and Royal Brisbane and Women’s Hospital (RBWH) HRECs (HREC/17/QRBW/616), and Uniting Care Health Human Research Ethics Committee (#1801), and the BMC dataset was approved by the University of Sydney HREC (2021/283). All participants provided written and informed consent to participate in research examining imaging biomarkers of Motor Neuron Disease and for deidentified data to be made available through publication and for other research purposes. To protect the identity of the participants, we constrained the field of view of the anatomical data to the mouth by cropping out data from elsewhere – i.e., no face information was included in the anatomical T1w images. Additionally, any identifiable information in the metadata has also been removed.

### Image acquisition

#### BeLong

Data from the BeLong study were collected between 2020-2023 and include 4 non-neurodegenerative healthy control (HC) participants. Control participants were recruited as a convenience sample of family, friends and colleagues of patients enrolled via the Motor Neuron Disease clinics at the Wesley Hospital and the RBWH. Imaging was performed at the University of Queensland, Centre for Advanced Imaging using a 3T Siemens Prisma (PrismaFit, Siemens Healthineers, Erlangen, Germany) using a 64-channel head and neck coil.

Participants were imaged using a 3D SPACE T2w sequence for spinal cord imaging covering the tongue with an isotropic resolution of 0.8mm^3^. This sequence shows high contrast of the fascia of the tongue and surrounding tissue and was acquired with the following parameters: TR=1500ms, TE=120ms, TA=4m:02s, FA=120°, matrix size=256×320, and number of slices=64.

#### EATT4MND

Participant information from the EATT4MND study has been described elsewhere^25^. Briefly, 24 HCs were imaged at the Herston Imaging Research Facility, Brisbane of which 19 participants are included in our data repository. Control participants were recruited as in the BeLong study. Data were collected using a 3T Siemens Prisma (Siemens Healthcare, Erlangen, Germany). T2-weighted (T2w) scans were obtained from a 3D SPACE 1mm^3^ isotropic sequence with the following parameters: TR = 5000ms, TE = 386ms, TI = 1800ms, TA = 5m:52s, matrix size =256×256 and number of slices=176^25^.

#### BMC

Whole-brain imaging was performed using a 3T MRI scanner (GE MR750, DV29; 32-channel Nova head coil) at the Brain and Mind Centre (BMC), The University of Sydney, Australia. Imaging data from 25 HCs were collected, 24 participants are included in our repository. Healthy control participants were recruited via study advertisement flyers and word-of-mouth. All healthy participants were screened for medical history. Written consent was provided by all participants prior to commencement of any research activities. Coronal T1-weighted images were acquired using an 1mm^3^ isotropic MPRAGE sequence (parameters: TE = 2.3ms, TR = 6.2ms; TI = 500ms, FA = 12°; matrix size = 256×256; number of slices = 204, TA = 5m 31s).

### Semi-automated tongue segmentation

#### Stage 1 – Active learning

To generate annotated data for segmentation model training, we initially manually annotated the BeLong dataset T2w images from healthy controls and MND patients. Specifically, three tongue volumes from three scans were manually annotated by XZ (Medical Principal House Officer with three years medical experience). These initially labelled data have been used in conjunction with MONAI Label^26^ within the Slicer application^27^ available on Neurodesk^28^ to interactively annotate the BeLong study data and iteratively train a model for T2w MRI tongue muscle segmentation. MONAI Label is based on active learning, which is a strategy that starts off by training a segmentation model on limited annotated data, which is then used for selecting a new sample from a pool of unlabelled data that may be labelled to improve model’s performance in the next iteration of model training. In this context, sample selection is based on the model’s uncertainty^29^. Using this framework, we trained a DynUNet model to segment five key tongue muscles: genioglossus, transverse and vertical muscles combined, superior longitudinal, and inferior longitudinal (Figure 1a), using default data augmentation strategies, including random flip and intensity scaling. In this iterative process, XZ, FLR (six years medical imaging experience), and TBS (ten years medical imaging experience) corrected and labelled 9 additional scans with assistance from XY.

**Figure 1.**
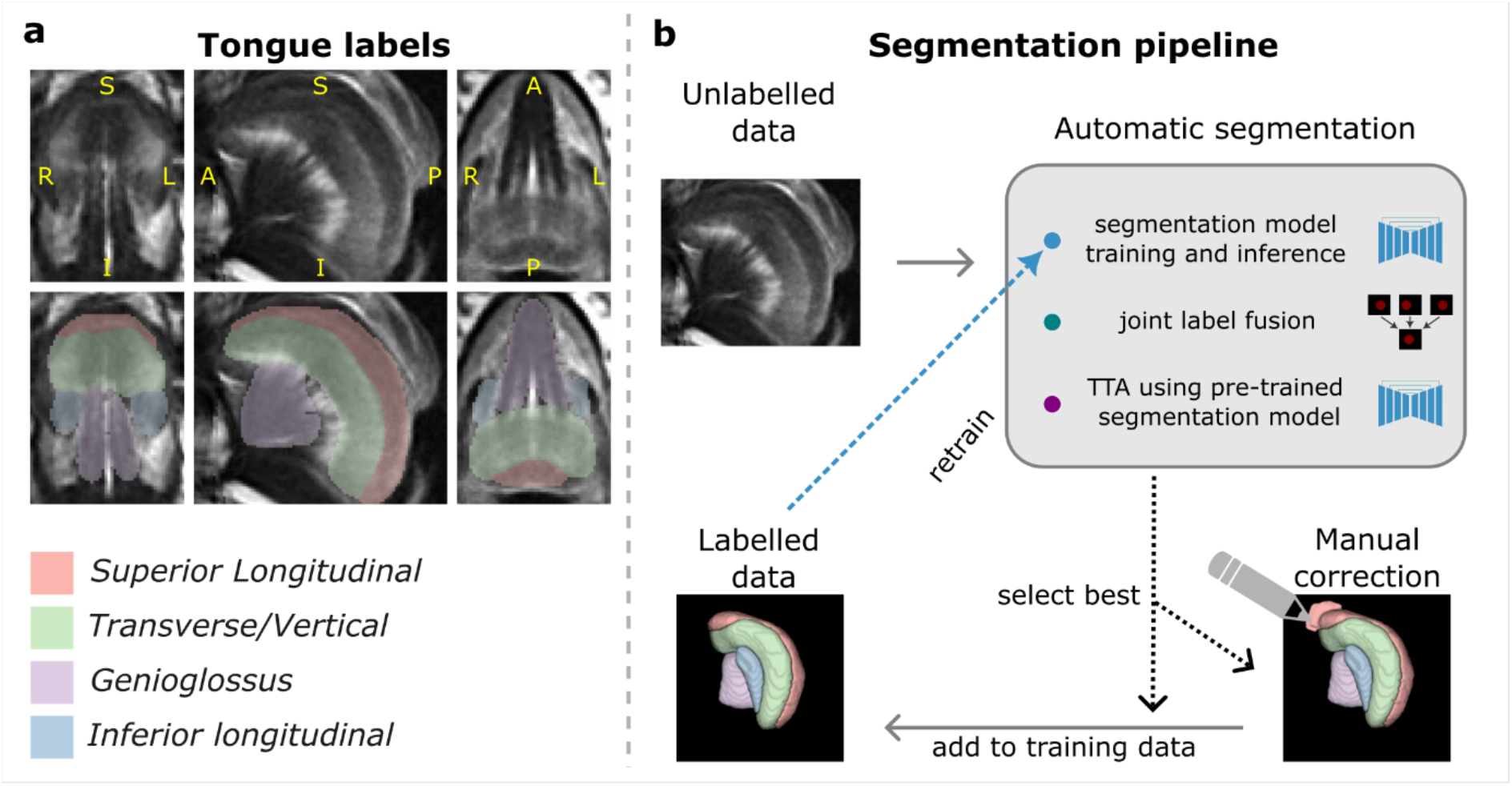
Tongue segmentation method. **a:** Atlas generated with Joint Label Fusion showing the labelled muscles and overlaid on the BeLong dataset template using T2w data. **b:** The segmentation pipeline: we gathered unlabelled data from each study and applied one of the three semi- or automatic segmentation methods to obtain rough segmentations. We then selected the best outputs by visual inspection, and either manually corrected the labels and/or added these data to our training for further segmentation model retraining. We then repeated this procedure until all data were labelled.

#### Stage 2 – Segmentation model training and inference on new datasets

After annotating and training on data from 12 individuals interactively and iteratively using MONAI Label, we used our own training implementation (Table 2) where data augmentation strategies were adjusted, and a new model was trained and used to predict tongue muscle segmentation on new unlabelled data. These initial predictions were further refined with manual correction (by FLR and TBS) and were leveraged with different approaches to speed up data annotation, including Joint Label Fusion (JLF)^30^ and test-time adaptation (TTA)^31,32^. We performed a few iterations of model training and prediction on unlabelled data whenever we had more refined segmentations available. This involved manually inspecting segmentations generated across all strategies and selecting the ones to be added to the training set in the next model training iteration (Figure 1b).

**Table 2.** Hyperparameters for segmentation model.

| Parameter name | Parameter value |
| --- | --- |
| Network architecture | DynUNet(*spatial_dims*=3,<br>*n_channels*=1,<br>*out_channels*=5,<br>*kernel_size*=[3, 3, 3, 3, 3, 3],<br>*strides*=[1, 2, 2, 2, 2, [2, 2, 1]],<br>*upsample_kernel_size*=[2, 2, 2, 2, [2, 2, 1]],<br>*norm_name*="instance",<br>*deep_supervision*=False,<br>*res_block*=True,<br>*dropout*=.2) |
| Optimizer | Adam |
| Learning rate | 0.0001 |
| Loss function | DiceCELoss |
| Number of epochs | 1000 (at each iteration of model training) |

##### Joint Label Fusion

JLF^30^ is a multi-atlas segmentation method that allows for combining a representative set of manually labelled datasets (or atlases) through data warping and weighted voting (label fusion). In detail, the technique involves non-linearly registering the atlases to the input image, then assigning a segmentation label to each voxel based on intensity similarities. Here, we used the 12 manually labelled data from Stage 1, which included both HC and patients’ data. We used the ANTs implementation of JLF^33,34^.The JLF atlas was used to estimate segmentation in unlabelled data through image registration or as a proxy (suboptimal segmentation) for segmentation model adaptation.

##### Test-time adaptation

TTA refers to a strategy for improving deep learning model generalisability to new data that follows a different distribution than the original training data^31,32^. For example, a model pre-trained on T2w MRI data from the BeLong study performs poorly on data from different collection sites (EATT and Sydney) or of different contrast weightings (i.e. T1w). Here, we used TTA to adapt our previously pre-trained model using a model adapter (a smaller convolutional neural network prepended to the segmentation model), aiming to improve the predicted segmentations for T1w data and data from different studies. The model adapter constituted of a small convolutional neural network with 3 layers (with up to 16 channels, kernel size = 3, and stride = 1) and an expressive activation function (act(x) = exp(−*x*^2^/*s*^2^); where *s* is a trainable parameter) applied to the output of the first and second layers, similar to previous work^35^. Note that only the parameters of the model adapter were trained while the segmentation model’s parameters were fixed to retain the segmentation knowledge learned from the BeLong dataset. Specifically, to guide model adaptation in a supervised fashion, we leverage a proxy (or suboptimal) segmentation that consists of the JLF atlas registered to each individual’s space. We performed instance-wise adaptation, i.e., the model adapter’s parameters were adapted (or trained) for each individual separately.

#### Stage 3 – Final manual correction

Each of these approaches were used to grow the pool of annotated and manually corrected data for a new iteration of segmentation model training. We iteratively “bootstrapped” the best segmentation out of the three methodologies (using a pre-trained model, TTA, and JLF) to manually correct the segmentation if required (Figure 1b). The new labelled data were then used to train a new segmentation model for the following iteration. We repeated this process until all data (HC and patient data) were segmented accurately, i.e., following segmentation landmarks according to our established method^20^. Our approach was intentionally multi-modal, as it was necessary to leverage the strengths of each method, given the variety of data (patient and control, different scanners, different MR contrasts). Finally, for the final release of HC data, FLR and TBS inspected, manually corrected, and smoothed all segmentations using a 1mm gaussian smoothing kernel across all labels independently.

### Data records

This dataset^21^ is deposited in the Open Science Framework (OSF), a free and open platform to support open research. The data can be accessed through this link: https://osf.io/wt9fc/ (DOI: 10.17605/OSF.IO/WT9FC). The files are organized per study, as shown in Figure 2.

**Figure 2.**
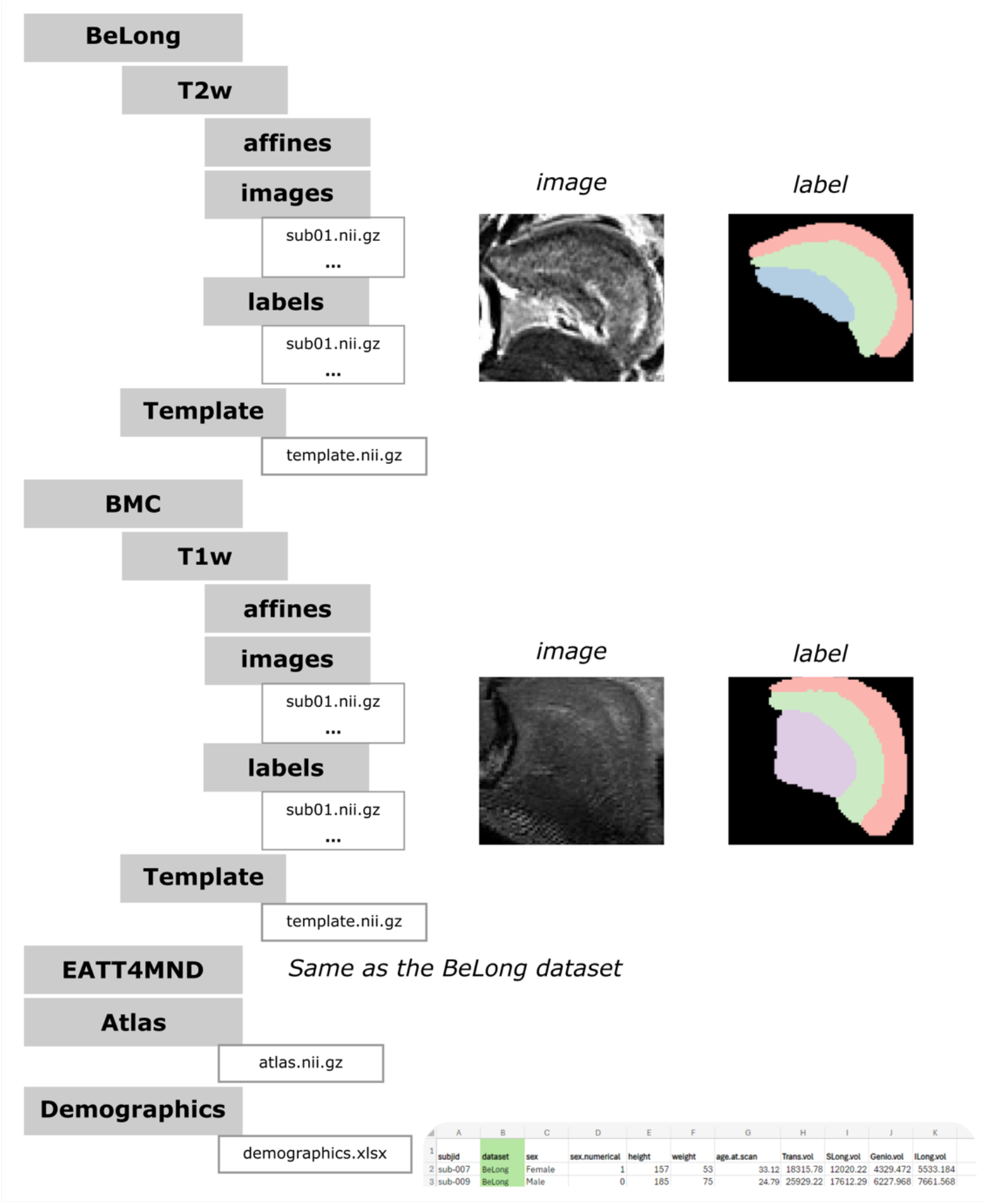
Summary of the dataset. The dataset is structured into distinct studies. For each study we provide either T1w or T2w images with corresponding segmentations, and a study template that can be used for new imaging data registration. We also provide an atlas generated with Joint Label Fusion and demographic information.

#### Imaging data

Under each study directory are folders for cropped T1w or T2w data. Within those folders, anatomical images are found in the <u>images</u> folder and corresponding segmentations are found in the <u>labels</u> folder. Note that this data has been registered to study templates generated as described below. Finally, to protect the identity of the participants, we constrained the field of view of the anatomical data to the mouth by cropping out data from elsewhere. The same transformation was applied to the labels. Figure 3 shows example data derived from each of these studies.

**Figure 3.**
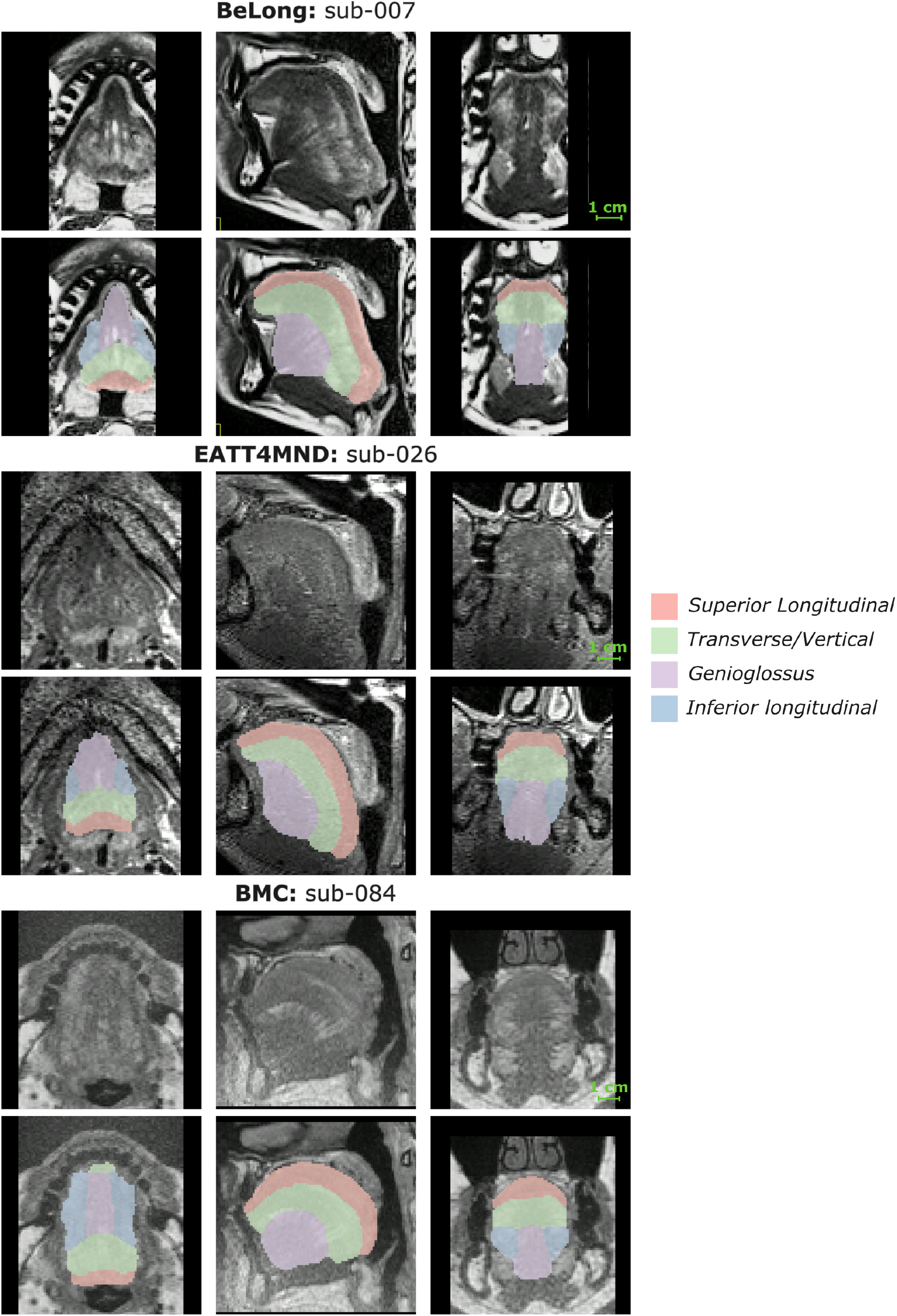
Image and label data examples for each study. An example image and label data are shown for each dataset (BeLong: top; EATT4MND: middle; BMC: bottom). The top row of each panel shows the MRI data (T2w images from BeLong and EATT4MND and T1w from BMC) and the bottom row shows the labels overlaid upon the MRI data.

Also under each study directory is a folder for a template that can be used for registration of new imaging data, if required. These templates were generated using data from both HC and patients using *antsMultivariateTemplateConstruction2*.*sh* from ANTs^33^. Parameters were: 3 iterations using an affine template, 8 iterations using a refined SyN template at 0.25 gradient step, and 6 more iterations using refined SyN template at 0.15 gradient step.

#### JLF atlas

We provide a JLF atlas generated with manually corrected segmentation from the BMC study, which can be found under the <u>Atlas</u> folder. The JLF atlas can be used to estimate segmentation in new unlabelled data through image registration, for example, using ANTs^33^ or the manual registration tool from ITK-SNAP^36^. Note that the provided atlas was generated with the final segmentations from the BMC study, i.e., it is not the same as the one described in the “Semi-automated tongue segmentation” section. The provided atlas was generated as described previously and with minimal manual correction of small mis-segmentations.

#### Demographic information

This dataset also includes demographic information (age, weight, height, and sex) and is available under the <u>Demographics</u> folder^21^.

## Technical validation

The quality of anatomical data has been evaluated by estimating signal-to-noise ratio (SNR) and contrast-to-noise ratio (CNR). SNR was calculated by first defining a noise region outside the brain and skull and dividing the mean intensity within the whole tongue musculature by the standard deviation of the noise region. CNR was calculated by subtracting the mean intensity value of the first tissue class (Transverse/Vertical) by the second tissue class (Superior Longitudinal), then dividing the result by the standard deviation of the noise region. All data processing was conducted using Nibabel^37^ in Python. Both measures are available in the demographic information spreadsheet and are represented in Figure 4a. Overall, the distribution of SNR was similar across datasets with observable trends of higher SNR in the EATT dataset and BeLong, although the latter has a smaller sample size (n=4). Additionally, the BeLong dataset has shown superior CNR compared to EATT and BMC datasets, which can also be observed in Figure 3. Nonetheless, despite observable differences in CNR across datasets, the distributions of total tongue volume (Figure 4a) and individual muscle’s volume (Figure 4b) were consistent across datasets, highlighting that both T1w and T2w MR contrasts offer enough contrast for tongue muscle segmentation.

**Figure 4.**
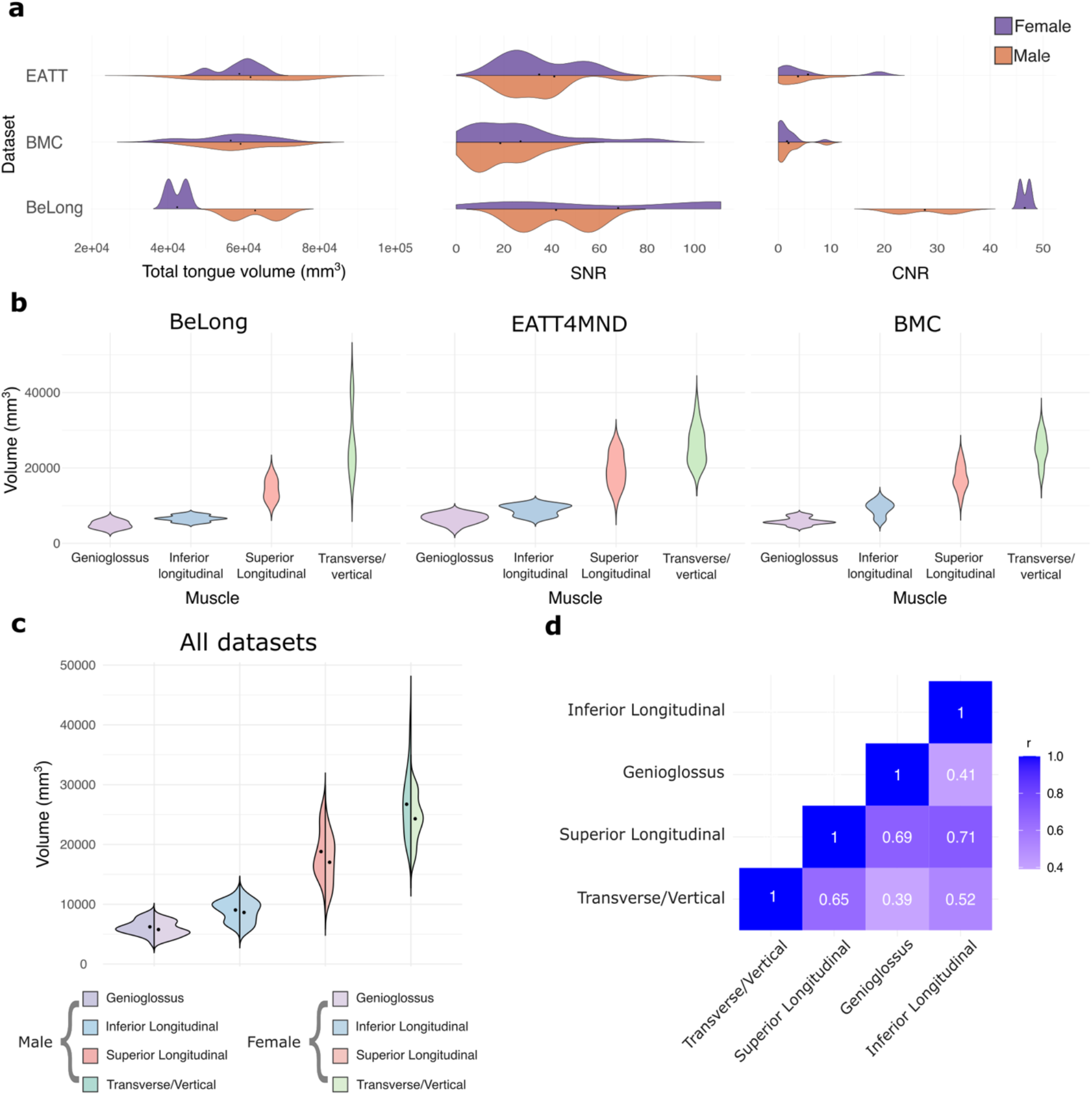
Data quality metric and muscle volume distributions. **a**, Distributions of total tongue volume (left), signal-to-noise ratio (middle), and contrast-to-noise ratio (right) across datasets. **b**, Distribution of tongue muscles’ volumes across datasets. **c**, Distributions of tongue muscles’ volumes between sex aggregated across datasets. **d**, Pair-wise correlation among inferior longitudinal, genioglossus, superior longitudinal, and transverse/vertical muscle volumes using all data.

With aggregated data from all datasets, we did not observe differences across sex (Figure 3c). To show the relationship between muscles (reflecting internal consistency of the muscle segmentations), we determined the correlation (Pearson’s r) between pairs of muscle’s volumes (Figure 3d), revealing different muscle volumes are generally well correlated with the exception of the genioglossus being less correlated with inferior longitudinal and transverse/vertical muscles.

## Code availability

All accompanying Python, R, and Bash source code is available on GitHub (https://github.com/thomshaw92/TongueSegMND).

## Data Availability

This dataset is deposited in the Open Science Framework (OSF), a free and open platform to support open research. The data can be accessed through this link: https://osf.io/wt9fc/.

## Acknowledgement

The authors acknowledge funding by a Motor Neuron Disease Research Australia (MNDRA) Postdoctoral Research Fellowship (PDF2112), NHMRC Ideas grant APP202987, FightMND Collaborative Initiative Grant, Lenity Australia, and an ARC Linkage grant (LP200301393). The data collection for EATT4MND was supported by funding from the Wesley Medical Research grant (#2017-07) and the University of Queensland, Faculty of Medicine. STN acknowledges funding from the Scott Sullivan MND Research Fellowship (MND and Me Foundation, RBWH, and the University of Queensland).

We thank all participants for their contributions. We also thank the radiology and professional staff at the Herston Imaging Research Facility, National Imaging Facility, UQ Centre for Clinical Research, and the UQ Research Computing Centre. We also thank Dr Diana Lucia for their support in collecting data. We acknowledge support from Aiman Al Najjar, Nicole Atcheson, Sarah Daniel, and the Centre for Advanced Imaging.

## Author contribution

**Fernanda L. Ribeiro:** Conceptualization, Methodology, Software, Validation, Formal analysis, Investigation, Data curation, Writing – Original Draft, Review & Editing, Supervision, Project administration, Visualization. **Xiangyun Zhu:** Methodology, Validation, Data curation, Writing – Review & Editing. **Xincheng Ye:** Methodology, Data curation, Writing – Review & Editing. **Sicong Tu:** Methodology, Resources, Data curation, Writing – Review & Editing, Funding acquisition. **Shyuan Ngo:** Resources, Writing – Review & Editing, Funding acquisition. **Robert Henderson:** Resources, Writing – Review & Editing, Funding acquisition. **Frederik Steyn:** Resources, Writing – Review & Editing, Funding acquisition. **Matthew Kiernan:** Funding acquisition. **Markus Barth:** Resources, Supervision, Writing – Review & Editing, Funding acquisition. **Steffen Bollmann:** Methodology, Writing – Review & Editing, Funding acquisition. **Thomas B. Shaw:** Conceptualization, Methodology, Software, Validation, Formal analysis, Investigation, Resources, Data curation, Writing – Original Draft, Review & Editing, Visualization, Supervision, Project administration, Funding acquisition.

